# Elenagen, a p62/SQSTM1-encoding plasmid, improves overall survival in platinum-resistant ovarian cancer patients: a phase II trial

**DOI:** 10.1101/2025.11.04.25339335

**Authors:** Sergei Krasny, Yauheni Baranau, Evgeny Bakin, Sergey Polyakov, Olga Streltsova, Ekaterina Zharkova, Aliona Filimonava, Gabriel Levin, Vladimir Gabai, Alexander Shneider

**Author notes:** These authors have contributed equally to this work (Sergei Krasny, Yauheni Baranau).

## Abstract

**Background:** Platinum-resistant ovarian cancer remains a major therapeutic challenge, with limited benefit from currently available cytotoxic agents. Elenagen is a newly developed plasmid DNA–based anticancer agent that encodes p62/SQSTM1 protein, a multifunctional adaptor protein involved in selective autophagy, signal transduction, and modulation of the inflammatory response. We previously reported the progression-free survival outcomes of patients with platinum-resistant ovarian cancer treated with Elenagen. We report the overall survival results from a phase II randomized controlled trial comparing Elenagen plus gemcitabine with gemcitabine alone.

**Methods:** This open-label, prospective, randomized, two-center study enrolled women with platinum resistant ovarian cancer. Patients were randomly assigned (1:1) to receive gemcitabine monotherapy (1,000 mg/m^2^ on days 1 and 8 every 21 days) or gemcitabine plus Elenagen (2.5 mg intramuscularly weekly). The primary endpoint was OS; secondary endpoints included safety, post-progression outcomes, and time-dependent analyses of Elenagen exposure. Survival was analyzed using Kaplan–Meier and Cox proportional hazards models.

**Results:** Thirty patients (15 per arm) were evaluable for overall survival. Baseline demographic and clinical characteristics were balanced between groups. Among patients with elevated CA-125 levels (>35 U/mL), median overall survival was 13 months (95% CI, 10–27) in the gemcitabine arm and 25 months (95% CI, 17–not reached) in the Elenagen plus gemcitabine arm (log-rank p = .031). Treatment with Elenagen was associated with a 59% reduction in the risk of death (HR = 0.41; 95% CI, 0.18–0.94; p = .036). Time-dependent and landmark analyses demonstrated a positive association between longer Elenagen exposure and improved survival (p<.001). No additional safety signals were observed compared with gemcitabine alone. Post-progression survival and subsequent therapy patterns were comparable between arms.

**Conclusions:** The addition of Elenagen to gemcitabine significantly prolonged overall survival in patients with platinum-resistant ovarian cancer without increasing toxicity.

**Trial registration:** NCT05979298, 2023-08-07

## Introduction

Approximately 20,000 new cases of ovarian cancer (OC) are diagnosed each year in the United States(1). High-grade serous ovarian cancer (HGSOC) the most common histologic subtype of OC, accounts for approximately 70% of all cases(2). It is typically diagnosed at an advanced stages and is associated with a 5-year overall survival rate of only 43%(3). Even following standard of care treatment, consisting of cytoreductive surgery and platinum-based chemotherapy with or without maintenance treatment, the majority of HGSOC patients experience disease recurrence and eventually develop platinum resistance (4). For platinum-resistant OC (PROC), four cytotoxic agents are most commonly used: pegylated liposomal doxorubicin (PLD), paclitaxel, gemcitabine, and topotecan, each demonstrating comparable overall response rates of approximately10% - 15%, progression-free survival (PFS) of 3 - 4 months, and overall survival (OS) of about 12 months. Recently, an antibody-drug conjugate targeting the folate receptor alpha (FRα) was approved for PROC, however its indication is limited to patients with high FRα expression(5).Therefore, there remains a significant unmet need in the management of PROC, and novel therapeutics approaches aimed at improving long-term outcomes are urgently required.

We have developed a proprietary biologic agent, the plasmid Elenagen (EG), which encodes the human p62 (SQSTM1) protein. This protein plays an essential role in tumor formation, progression, metastasis, and the development of chemo- and immune-resistance (6-8). In preclinical studies, EG demonstrated broad anti-tumor and anti-metastatic activity in rodent tumor models (9), as well as in spontaneous canine mammary tumors (10) (11). It also demonstrated a favorable safety profile and preliminary evidence of efficacy in phase I clinical trial involving patients with various solid tumors (12). In multiple animal models of diseases linked to chronic inflammation, EG demonstrated systematic anti-inflammatory effects by reducing pro-inflammatory and elevating anti-inflammatory cytokines as well as modulating mesenchymal stem cells(13-17).

Recently, we conducted a prospective, randomized, multicenter study involving two cohorts of 20 patients each with PROC (18). We evaluated the addition of EG to single agent gemcitabine. Here, we report a sub analysis, which examines overall survival in patients enrolled in our study with CA-125 level greater than 35 U/mL.

## Materials and Methods

This was an open-label, prospective, randomized, two-center, two-arm study conducted between January 2020 and August 2024.

### Patients

#### Inclusion criteria

1. Female patients aged 18–70 years at the time of consent.
2. Histologically confirmed diagnosis of PROC.
3. Measurable disease according to RECIST 1.1 criteria.
4. PROC defined as recurrence within 6 months of last dose of platinum-based chemotherapy.
5. Provision of written informed consent.
6. Eastern Cooperative Oncology Group (ECOG) performance status of 0 or 1.
7. Estimated life expectancy of at least 6 months.
8. Adequate hematologic, renal, and hepatic function as specified by protocol defined laboratory thresholds.

#### Exclusion criteria

1. Presence of serious concomitant diseases or medical conditions that, in the investigator’s judgment, could interfere with study participation or protocol compliance.
2. Major surgery performed within 4 weeks prior to enrolment.
3. Radiotherapy involving an extended-field radiation within 4 weeks, or a limited-field within 2 weeks before study treatment initiation.
4. Administration of an investigational drug within 14 days or five half-lives (whichever is shorter) prior to the first study dose; or prior treatment with a monoclonal antibody within 30 days of the first study dose.
5. Known immediate or delayed hypersensitivity or idiosyncratic reaction to agents chemically related to Elenagen.
6. Documented intolerance or allergy to the study drug or any of its components.
7. Prior exposure to the same chemotherapy regimen as that assigned to the patient’s study cohort.

### Study design

Patients were randomized in a 1:1 ratio into two treatment cohorts, comprising twenty patients each. In the gemcitabine monotherapy group, patients received gemcitabine at a dose of 1,000 mg/m^2^ on days 1 and 8 of a 21-day cycle. In the EG combination group, patients received the same gemcitabine regimen supplemented with EG (2.5 mg administrated intramuscularly once weekly). Per protocol, upon disease progression, patients were permitted to transition to alternative standard chemotherapeutic regimens, which continued for as long as clinical benefit was observed and the patient remained alive. EG treatment was to be maintained despite disease progression, until patient withdrawal of consent (which did not occur in any case) or death. Because of force majeure, EG administration was simultaneously stopped for all patients, regardless of their prior treatment duration.

### Outcomes

In our previous report, we reported PFS assessed according to RESIST v1.1 criteria and safety evaluated using the National Cancer Institute Common Terminology Criteria for Adverse Events (NCI CTCAE version 5.0) (18). In this sub analysis, the primary end point was OS, defined as the time from randomization to death from any cause. Our secondary endpoints were PFS 2, and time dependent EG efficacy.

### Statistical analysis

Categorical variables were summarized as frequencies and percentages, while continuous variables were expressed as medians and interquartile range (IQR). In the EG arm, treatment duration varied among patients, therefore, a Cox proportional hazards regression analysis was performed with EG treatment duration included as a time-dependent covariate.

The analysis was complemented by a landmark approach, in which the total duration of pre-landmark treatment was included as a baseline covariate. This method allows assessment of the time-dependent effect of treatment within a defined observation window (19, 20). In a landmark plot, the X-axis represents distinct time points (“landmarks”) following treatment initiation, while the Y-axis depicts the logarithm of the hazard ratio (HR) comparing patients who had received treatment for at least that duration with those who had received it for a shorter duration or not at all, as estimated by the Cox proportional hazards model. A negative value at a given landmark indicates that treatment duration equal to or exceeding that time point is associated with a lower subsequent risk of death. Conversely, as the value approaches zero, it suggests that beyond that time point, no statistically significant survival benefit from continued treatment can be detected.

Multiple landmark time points were evaluated in order to characterize the temporal dynamics of the EG treatment throughout the course of the trial. All reported confidence intervals were 95% while p < .05 was defined the significance threshold. Analysis was performed by the R software, (version 4.5.1; R Foundation for Statistical Computing, Vienna, Austria)

## Results

### Patient characteristics

The data cutoff occurred 24 months after completion of treatment for the last enrolled patient, with a total study duration of 56 months. Patient characteristics are shown in **Table 1**. A total of thirty patients were included in the OS analysis, with fifteen in the control group and fifteen in the EG group. Baseline demographic and clinical characteristics were balanced between arms. The median age was 56 years [IQR 52-63] in the control group and 54 [IQR 47-62] in the EG group (p = .389). Serous histology was the predominant subtype (87% in both groups). Clear cell carcinoma was observed in 13% of controls and 6.7% of EG patients. Most tumors were grade 3 (79% vs. 100%, p = .389). The number of prior platinum-based therapy lines was comparable between study groups (p = .662), with the majority having received one prior line. Baseline CA-125 levels were similar in both study groups median 132 [IQR 93-525] vs. 101 [IQR 68-485] (p = .539). Peritoneal metastases were present in 87% of controls and 93% of EG patients (p > .99). Ascites was observed in 60% and 47% of patients, respectively (p = .464).

**Table 1.**
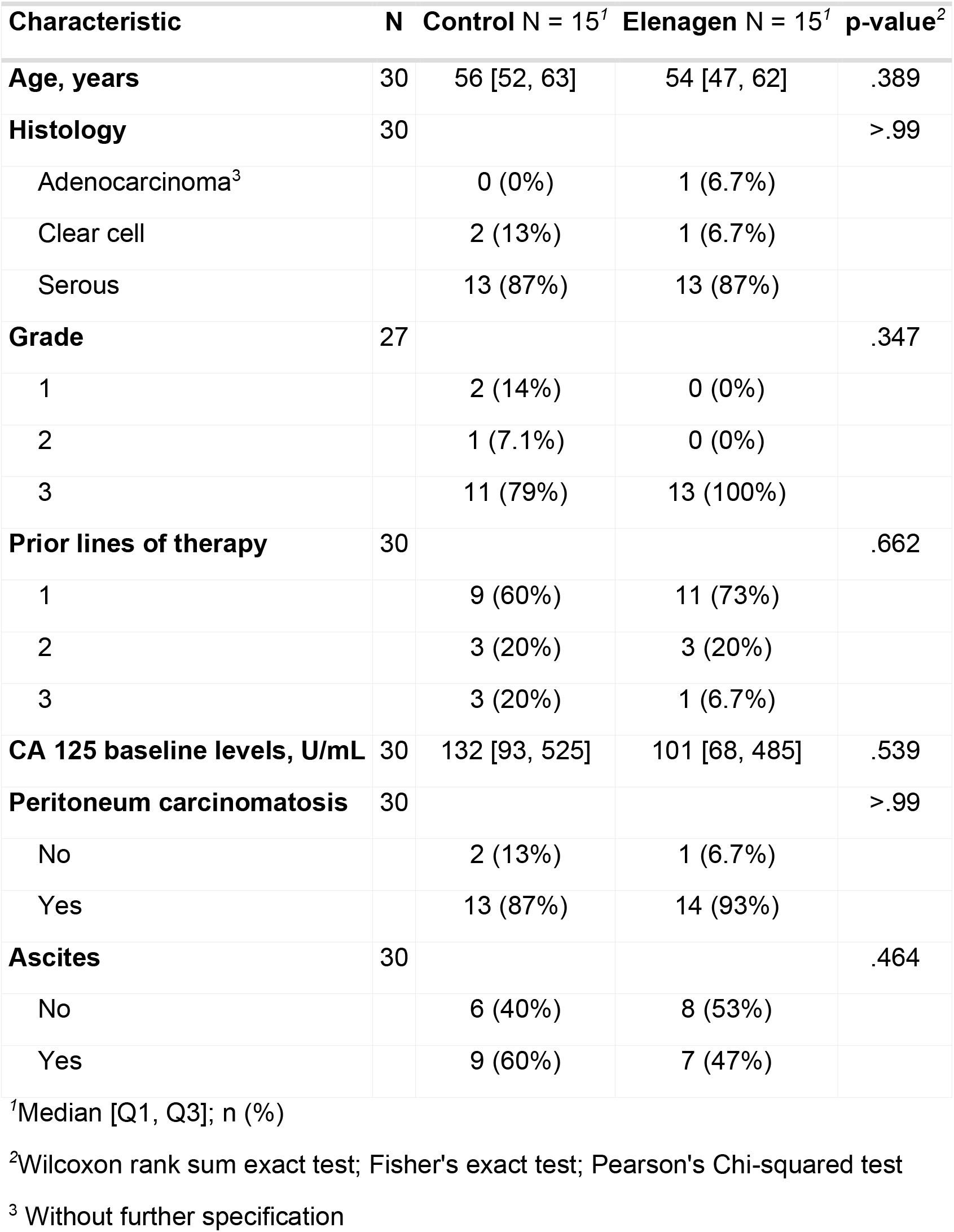
Patients’ characteristics.

| **Characteristic** | **N** | **Control N = 15<sup>1</sup>** | **Elenagen N = 15<sup>1</sup>** | **p-value<sup>2</sup>** |
| --- | --- | --- | --- | --- |
| **Age, years** | 30 | 56 [52, 63] | 54 [47, 62] | .389 |
| **Histology** | 30 |  |  | >.99 |
| Adenocarcinoma<sup>3</sup> |  | 0 (0%) | 1 (6.7%) |  |
| Clear cell |  | 2 (13%) | 1 (6.7%) |  |
| Serous |  | 13 (87%) | 13 (87%) |  |
| **Grade** | 27 |  |  | .347 |
| 1 |  | 2 (14%) | 0 (0%) |  |
| 2 |  | 1 (7.1%) | 0 (0%) |  |
| 3 |  | 11 (79%) | 13 (100%) |  |
| **Prior lines of therapy** | 30 |  |  | .662 |
| 1 |  | 9 (60%) | 11 (73%) |  |
| 2 |  | 3 (20%) | 3 (20%) |  |
| 3 |  | 3 (20%) | 1 (6.7%) |  |
| **CA 125 baseline levels, U/mL** | 30 | 132 [93, 525] | 101 [68, 485] | .539 |
| **Peritoneum carcinomatosis** | 30 |  |  | >.99 |
| No |  | 2 (13%) | 1 (6.7%) |  |
| Yes |  | 13 (87%) | 14 (93%) |  |
| **Ascites** | 30 |  |  | .464 |
| No |  | 6 (40%) | 8 (53%) |  |
| Yes |  | 9 (60%) | 7 (47%) |  |

<sup>1</sup>Median [Q1, Q3]; n (%)<sup>2</sup>Wilcoxon rank sum exact test; Fisher's exact test; Pearson's Chi-squared test<sup>3</sup> Without further specification

### Effect of EG on overall survival

Among patients with elevated CA-125 levels (>35 U/ml) which comprised the majority in both cohorts (15 of 20 patients) Kaplan–Meier analysis (**Fig.1**) revealed a median OS of 13 months (95% CI 10-27) in the gemcitabine group, vs. a median OS of 25 months (95% CI 17-not reached) in EG + gemcitabine arm (p = .031, log-rank), with approximately one-quarter of patients in the EG + gemcitabine cohort surviving beyond 48 months (**Fig.1)**. In Cox proportional hazard model, EG + gemcitabine treatment was associated with a reduction of the risk of death with a hazard ratio of 0.41 (95% CI 0.18-0.94, p= 0.036) (**Table 2)**.

**Figure 1.**
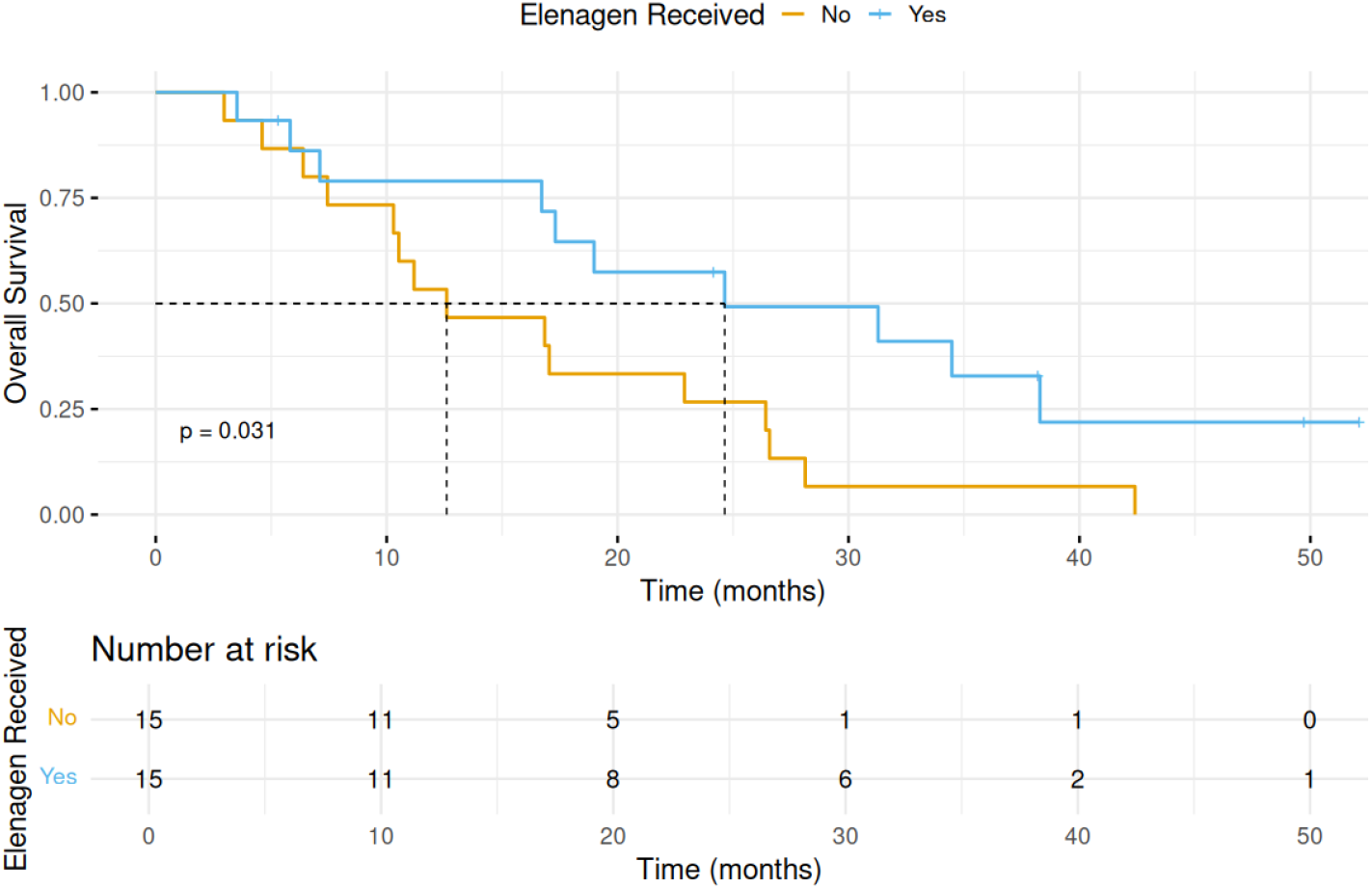
Overall survival in gemcitabine vs. EG + gemcitabine cohorts.

**Table 2.**
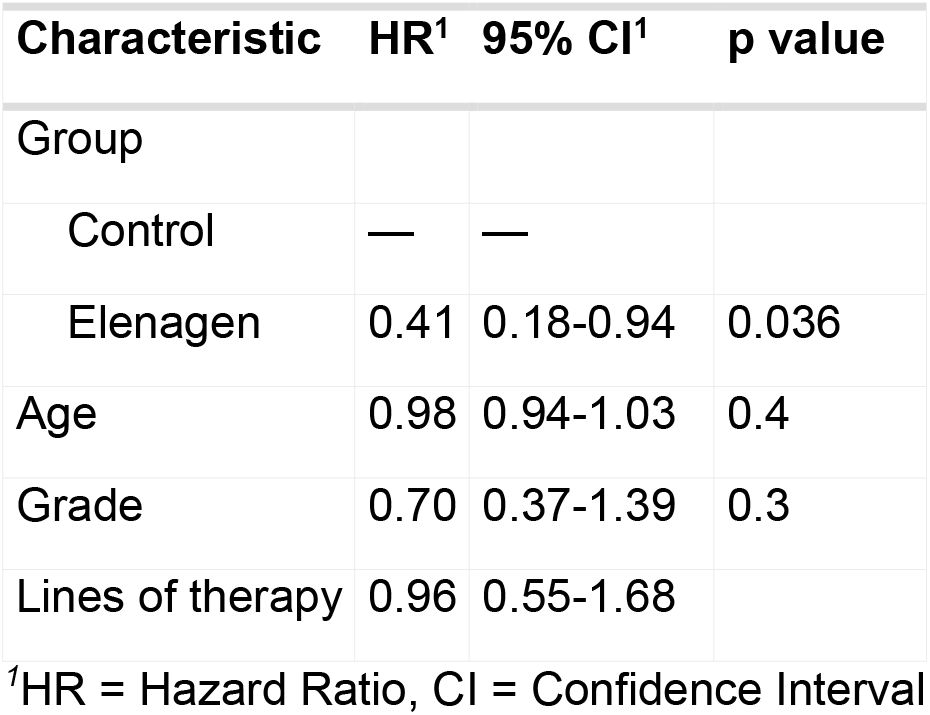
Cox proportional hazard model.

| Characteristic | HR<sup>1</sup> | 95% CI<sup>1</sup> | p value |
| --- | --- | --- | --- |
| Group |  |  |  |
| Control | — | — |  |
| Elenagen | 0.41 | 0.18-0.94 | 0.036 |
| Age | 0.98 | 0.94-1.03 | 0.4 |
| Grade | 0.70 | 0.37-1.39 | 0.3 |
| Lines of therapy | 0.96 | 0.55-1.68 |  |

<sup>1</sup>HR = Hazard Ratio, CI = Confidence Interval

### Dose-dependent effect of Elenagen treatment

The length of EG treatment depended on each patient’s enrolment date, ranging from 0.7 to 30.8 months, with a median of 10.2 months. In these patients, no additional signals of toxicity were noticed beyond our previous report. In a time-dependent Cox model - longer treatment with EG was associated with improved survival (p<.001) (**Table 3**).

**Table 3.**
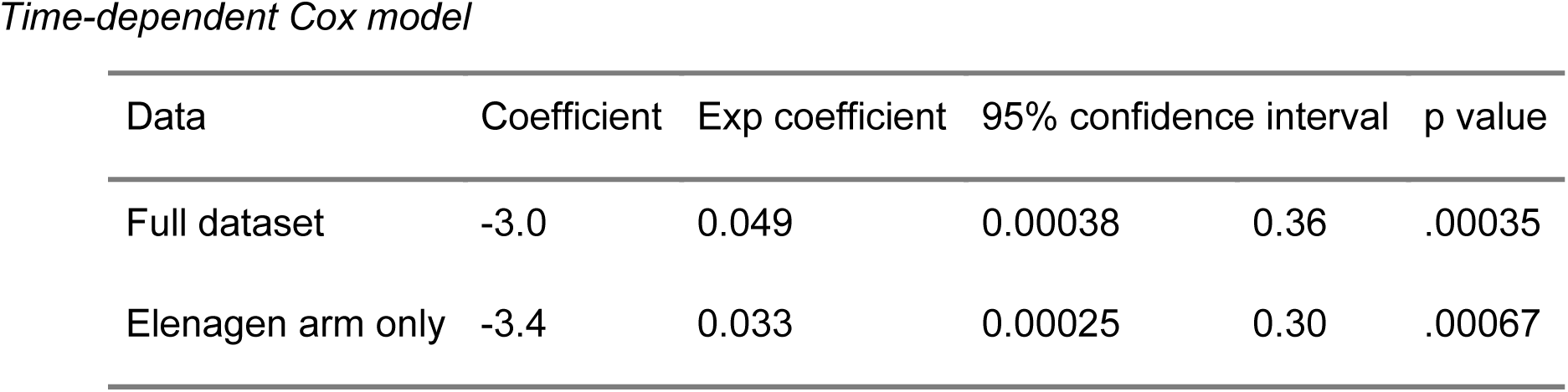
Analysis of Elenagen impact with time-dependent Cox models.

| Data | Coefficient | Exp coefficient | 95% confidence interval |  | p value |
| --- | --- | --- | --- | --- | --- |
| Full dataset | -3.0 | 0.049 | 0.00038 | 0.36 | .00035 |
| Elenagen arm only | -3.4 | 0.033 | 0.00025 | 0.30 | .00067 |

According to landmark analysis - longer EG treatment was associated with improved OS in GEM-treated PROC patients (**Fig 2**), with the strongest association observed during the first twelve months of therapy and remaining statistically significant thereafter (**Fig. 2, Table S1**).

**Figure 2.**
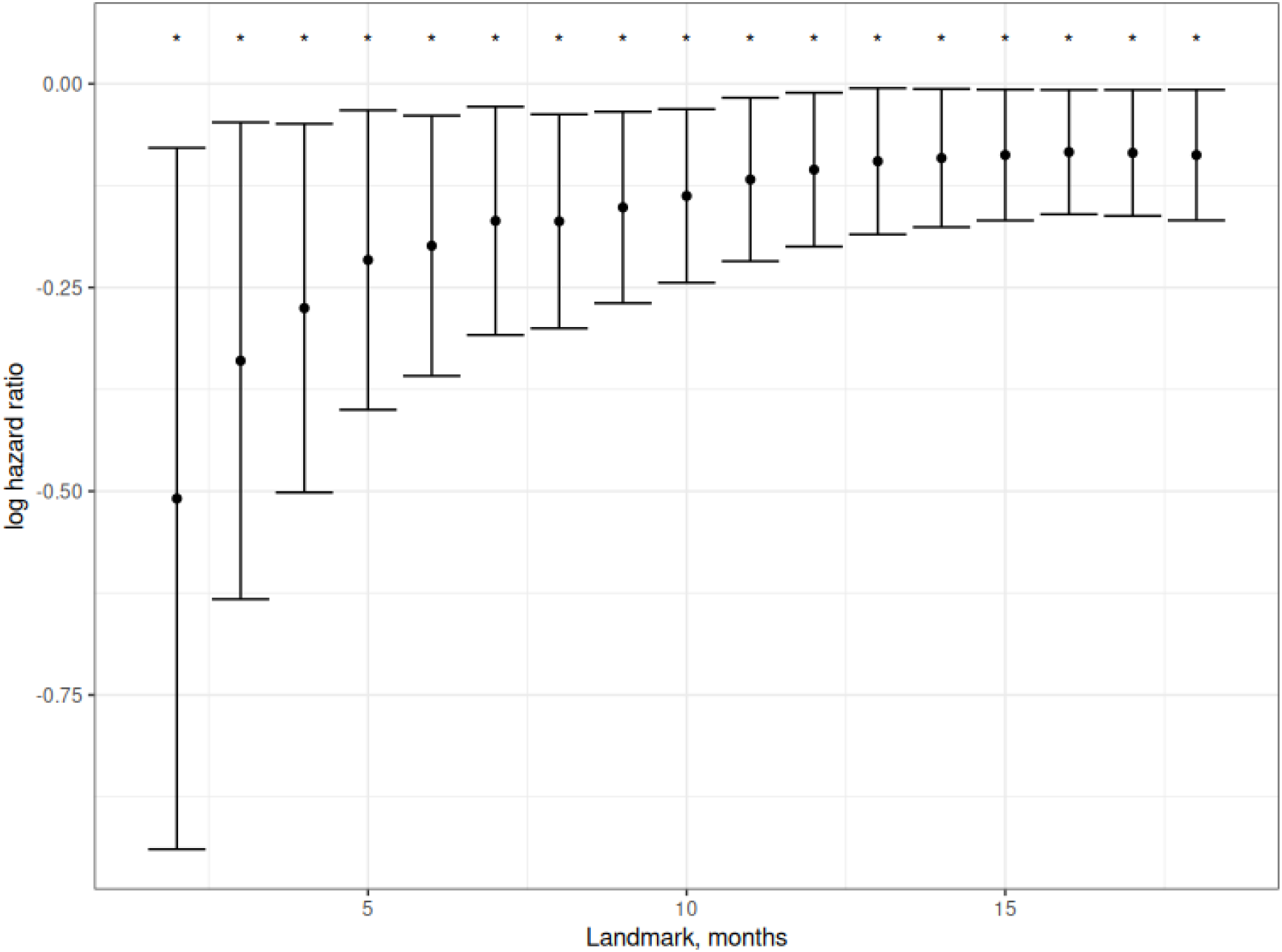
Landmark analysis: Post-hoc analysis with landmark method.

### Post Progression treatment

Following progression, all 19 patients in gemcitabine group and all 15 progressed patients in EG+ gemcitabine arm received further anticancer therapy. Various chemotherapy regimens were administered to all nineteen patients of gemcitabine group; in EG group, one patient underwent surgery and another – radiotherapy, while the remaining thirteen patients received chemotherapy regimens comparable to those used in the gemcitabine group (**Table S2**). In both arms the most common drugs used were taxanes and irinotecan, and the medium number of lines of chemotherapy was 3 (range 1-7) (Sup. Table 2). At the time of data cutoff, all patients who had received subsequent therapy had experienced disease progression. There was no difference in PFS 2 between EG and gemcitabine arms with medium of about 6 months in both arms (**Fig. S1**).

## Discussion

### Summary of main results

In this OS analysis we report that the addition of EG to gemcitabine resulted in improved OS of 12 months.

### Results in the Context of Published Literature

PROC has a dismal prognosis with OS of approximately12 months (21, 22), and novel therapy with antibody drug conjugates is designated for only biomarker driven indication(5). Previously, we demonstrated that the addition of our novel plasmid drug, Elenagen, to standard gemcitabine chemotherapy in patients with ROC significantly improved PFS, extending it from 2.8 months to 7.2 months. Importantly, no evidence of increased toxicity was observed with the combination compared to gemcitabine monotherapy. (18).

In a recent trial of relacorilant, a first-in-class selective glucocorticoid receptor antagonist, in PROC patients, its addition to nab-paclitaxel therapy increased OS from 11·50 months to 15.97 months (23). Antibody-drug conjugates (ADCs) have emerged as a promising class of anticancer drugs, delivering cytotoxic agents directly to tumor cells while sparing normal tissue. The FDA recently approved mirvetuximab soravtansine (MIRV) targeting folate receptor-positive PROC which demonstrated improved PFS (5.62 vs. 3.98 months) and overall survival (16.46 vs. 12.75 months). Importantly, although cross trial comparisons are questionable, specifically when different agents are discussed, our results demonstrate a higher benefit with EG. Additionally, adverse effects with MIRV such as blurred vision, nausea, diarrhea, and fatigue are common, though most are mild to moderate (24) (25). Furthermore, in contrast to MIRV, which is effective only in patients with high folate receptor alpha expression, thereby limiting its applicability to approximately 30–35% of PROC cases, Elenagen demonstrated benefit irrespective of biomarker status (26) (27).

NF-κB pathway, is a major pathway of inflammatory signaling (28, 29). OC with activated NF-κB signaling, are generally associated with poor prognosis owing to the upregulation of genes that promote malignant progression as well as immune and chemoresistance e (29).

We hypothesize that the sensitizing effect of EG on chemotherapy with gemcitabine in PROC can be related, in particular, to its inhibitory effect on NF-kB signaling. First, it was shown that gemcitabine can activate NF-kB and thus confer resistance to gemcitabine via feed forward loop (30), (31) (32). Second, inhibition NF-kB signaling sensitizes to gemcitabine both cancer cells in vitro and tumors in vivo (31) (33) (34) (35). Finally, we found EG can inactivate NF-kB signaling and suppress its target cytokines (36, 37) breaking this feed forward loop. Therefore, we hypothesize that EG increases antitumor effect of gemcitabine in our trial, at least partially, via inactivating NF-kB signaling. If our hypothesis is correct, we may expect that potentiation of gemcitabine therapy by EG may be observed in other cancers as well. Besides NF-kB signaling, inflammatory cytokines blocked by EG such as IL-1b, IL-6 and TNF also play a role in OC progression and chemo-immuno-resistance (38).

### Strengths and Weaknesses

The strength of this study lies in being the first to demonstrate a robust positive effect of the novel plasmid drug Elenagen on overall survival in advanced PROC, without visible side effects.

Noticeably, the main limitation is the small sample size (30 patients), underscoring the need for confirmation in a larger, multicenter population. In addition, the study was conducted at only two centers, potentially introducing institutional or geographic biases in patient selection and management practices. The open-label design may also have introduced bias in treatment administration or assessment of secondary endpoints, despite the objective nature of survival outcomes.

Furthermore, the study did not incorporate molecular stratification as *BRCA*, which could help identify patient subgroups most likely to benefit from EG. Finally, although the observed survival benefit is encouraging, longer follow-up and validation in larger, randomized, multicenter phase III trials will be essential to confirm the clinical efficacy and safety of EG in broader populations.

### Implications for Practice and Future Research

The results of this phase II randomized trial could offer a novel, well-tolerated treatment option for a population with limited therapeutic alternatives and poor prognosis. The observed dose-dependent survival effect highlights the biological activity of Elenagen and supports continued investigation of its immunomodulatory and NF-κB-inhibitory mechanisms. Future research should validate these findings in larger, multicenter phase III trials, incorporate molecular stratification, and explore the potential of Elenagen across other gemcitabine-sensitive solid tumors. Integration of biomarker-guided translational studies may also clarify the pathways through which Elenagen enhances chemosensitivity and contributes to durable disease control.

## Conclusion

The addition of EG to gemcitabine chemotherapy is effective in patients with PROC increasing OS without EG-associated side effects. Future studies of EG with various tumors and chemotherapy regimens, especially gemcitabine, are warranted.

**Contributors SK**: data curation, methodology, investigation, writing—original draft, writing—review, and editing. **AS**: Guarantor, data curation, methodology, investigation, writing—original draft, writing—review, and editing. **GL**: writing – original draft, data curation, methodology, formal analysis, writing—original draft, writing—review, and editing. **YB**: data curation, investigation. **EB**: methodology, writing – review and editing. **SP** - writing—review and editing. **OS**: writing—review and editing. **EZ**: writing— review and editing. **AF** – methodology, writing— review and editing. **VG** - conceptualization, methodology, project administration, writing—original draft, writing—review, and editing.

## Data Availability

All data produced in the present study are available upon reasonable request to the authors

## Patient consent for publication

Not applicable.

## Acknowledgments

We thank the patients and their families.

## Ethical approval

The study was approved by the Ministry of Health of Belarus (#03-19) and ethical review boards of N. Alexandrov National Cancer Centre of Belarus and Minsk City Cancer Center. Informed consents were signed by all study participants.

