## Supplementary material for "Elenagen, a p62/SQSTM1-encoding plasmid, improves overall survival in platinum-resistant ovarian cancer patients: a phase II trial": Suppl Tables 1, 2

Suppl **Table 1**

**Landmark analysis of effect of duration of treatment with Elenagen**

| Landmark_time | n | estimate | std.error | statistic | p.value | 2.5 % | 97.5 % |
|---|---|---|---|---|---|---|---|
| 2 | 30 | -0.510 | 0.220 | -2.3 | 0.020 | -0.94 | -0.0790 |
| 3 | 29 | -0.340 | 0.150 | -2.3 | 0.023 | -0.63 | -0.0470 |
| 4 | 28 | -0.280 | 0.120 | -2.4 | 0.017 | -0.50 | -0.0490 |
| 5 | 27 | -0.220 | 0.094 | -2.3 | 0.021 | -0.40 | -0.0330 |
| 6 | 25 | -0.200 | 0.081 | -2.4 | 0.015 | -0.36 | -0.0390 |
| 7 | 24 | -0.170 | 0.071 | -2.4 | 0.019 | -0.31 | -0.0280 |
| 8 | 22 | -0.170 | 0.067 | -2.5 | 0.012 | -0.30 | -0.0380 |
| 9 | 22 | -0.150 | 0.060 | -2.5 | 0.011 | -0.27 | -0.0340 |
| 10 | 22 | -0.140 | 0.054 | -2.5 | 0.011 | -0.24 | -0.0310 |
| 11 | 20 | -0.120 | 0.051 | -2.3 | 0.022 | -0.22 | -0.0170 |
| 12 | 19 | -0.110 | 0.048 | -2.2 | 0.029 | -0.20 | -0.0110 |
| 13 | 18 | -0.095 | 0.046 | -2.1 | 0.038 | -0.18 | -0.0054 |
| 14 | 18 | -0.091 | 0.043 | -2.1 | 0.035 | -0.18 | -0.0065 |
| 15 | 18 | -0.087 | 0.041 | -2.1 | 0.033 | -0.17 | -0.0071 |
| 16 | 18 | -0.084 | 0.039 | -2.2 | 0.031 | -0.16 | -0.0075 |
| 17 | 16 | -0.085 | 0.039 | -2.1 | 0.032 | -0.16 | -0.0075 |

| Landmark_time | n | estimate | std.error | statistic | p.value | 2.5 % | 97.5 % |
|---|---|---|---|---|---|---|---|
| 18 | 14 | -0.087 | 0.041 | -2.1 | 0.032 | -0.17 | -0.0074 |

Suppl **Table 2**

Post- progression treatments in Control (GEM only) and EG cohorts

| Subsequent anticancer Therapy | Elenagen | | Control | |
|---|---|---|---|---|
| Any | 15 | | 19 | |
| Surgery | 1* | | 0 | |
| Radiotherapy | 1 | | 3 | |
| Chemotherapy | 13 | | 19 | |
| Line's number | Range 1-7<br>Median 3 | | Range 1-7<br>Median 3 | |
| Taxanes (paclitaxel-weekly) | 11 | 84.6% | 12 | 63.2% |
| Irinotecan | 8 | 61.5% | 10 | 52.6% |
| Platinum (Cisplatin/Carboplatin) | 7 | 53.8% | 3 | 15.8% |
| Vinorelbine | 6 | 46.2% | 7 | 36.8% |
| Treosulfan | 6 | 46.2% | 9 | 47.4% |
| Capecitabine/Fluorouracil | 6 | 46.2% | 8 | 42.1% |
| Bevacizumab with Chemo | 6 | 46.2% | 7 | 36.8% |
| Oxaliplatin | 5 | 38.5% | 6 | 31.6% |
| Doxorubicin | 5 | 38.5% | 2 | 10.5% |
| ifosfamide | 3 | 23.1% | 2 | 10.5% |
| Etoposide | 2 | 15.4% | 2 | 10.5% |
| Methotrexate | 1 | 7.7% | 0 | 0.0% |
| Gemcitabine | 1 | 7.7% | 0 | 0.0% |
| Cyclophosphamide | 1 | 7.7% | 2 | 10.5% |
| Hormonotherapy (AI, SERM)) | 1 | 7.7% | 2 | 10.5% |
| Pemetrexed | 0 | 0.0% | 2 | 10.5% |
| TKI (MEK inhibitors) | 0 | 0.0% | 1 | 5.3% |

* Optimal cytoreduction after registered partial response in protocol treatment, remission ongoing.
